# Enhancing Stroke Awareness and Activation among High-Risk Populations: A Randomized Direct Mail Intervention in Diverse Healthcare Settings

**DOI:** 10.1101/2024.08.09.24311771

**Authors:** Christine C. Groves, Teresa M. Damush, Laura J. Myers, Fitsum Baye, Joanne K. Daggy, Holly Martin, Layne Mounsey, Daniel O. Clark, Linda S. Williams

**Author notes:** **Corresponding Author:** Christine C. Groves,. Indiana University School of Medicine, Department of Physical Medicine & Rehabilitation, 355 W 15^th^ St, Suite 3800, Indianapolis, IN 46202.

## Abstract

**Background:** Many patients are unaware of their stroke risk. The purpose of this research was to compare the effect of behaviorally tailored mailed messages on patient activation to reduce stroke risk.

**Methods:** We used electronic health records to construct Framingham Stroke Risk Scores (FSRS) in primary care patients from one Veterans Health Administration (VA) and one non-VA healthcare system, Eskenazi Health System (EHS). Four stroke risk messages were developed through patient interviews: standard, incentive ($5 gift card), salience, and incentive plus salience. Patients in the highest FSRS quintile were randomly assigned to receive one of the messages. All letters asked the patient to call a stroke prevention coordinator. Response to the messages was modeled separately in the two cohorts using logistic regression.

**Results:** From 6,695 eligible patients, 2,084 EHS patients (mean age 65.6, 36% male, 68% Black, mean FSRS 13.1) and 1,759 VA patients (mean age 75.6, 99% male, 86% White, mean FSRS 18.6) received a letter. Rates of calls to the coordinator were 13% among the EHS and 23% among the VA cohort. The EHS cohort was significantly more likely to respond to the incentive message compared to the standard message (OR = 1.97 [1.17, 3.09]), and the VA cohort was more likely to respond to the incentive plus salience message (OR = 1.50 [1.02, 2.22]). Older age (for VA) and Black race (for EHS) were also significantly associated with response. Among individuals calling the coordinator, 30% of the EHS cohort and 26% of the VA cohort were unaware they had stroke risk factors.

**Conclusions:** A mailed message including a $5 incentive was more effective than a standard message in engaging high-risk patients with their healthcare system; including a salience message may also be important in some patient populations. Many primary care patients are unaware of their stroke risk.

## Introduction

Stroke is a common problem that disproportionately impacts historically minoritized communities. Nearly 800,000 Americans have a stroke each year, and data suggest that although stroke incidence and mortality rates are declining overall, these declines are not as significant among individuals who are Black and Hispanic/Latina/Latino/Latinx. ^1^ Stroke prevalence and deaths are rising disproportionally among Black and Hispanic/Latina/Latino/Latinx individuals – a trend expected to continue.^1^ Socio-economic status (SES) is also strongly related to stroke and stroke risk factors, with lower SES associated with increased prevalence of diabetes and hypertension in the US.^1,2^ Even in the UK, with its national healthcare system, lower SES is associated with increased risk of stroke and worse post-stroke outcomes.^3^

Stroke risk factors are common, well described, and well recognized by the medical community, with established interventions available to reduce stroke risk. In fact, the continued decline of stroke incidence and mortality over the past several decades is a testament to the implementation and effectiveness of stroke prevention efforts across the US.^4^ Reasons why these gains are not realized among historically minoritized groups as well as those with lower SES are complex. The growing body of equity-focused research has uncovered many social determinants of health factors including income, area deprivation, education level, health insurance, social isolation, and access to high-quality, equitable healthcare.^5,6^ At the healthcare system level, it is imperative to improve access to effective stroke risk reduction interventions and programs that are readily available in the healthcare systems and communities they serve.^5–9^ Fundamental to this linkage of patient to healthcare system is activating at-risk patients to seek medical care and resources to change behavioral risk factors.

In this project, we drew on the science that has provided a foundation for behavioral economics as a guide to activation interventions. Specifically, we used the MINDSPACE framework that was created by some of the founders of behavioral economics.^10^ That framework details nine approaches to behavioral activation that have particularly solid evidence bases; we chose two of these approaches, salience, and incentives, based on their applicability to direct mail messaging strategies. Salience refers to things that are particularly relevant and noticeable to the individual. Incentives are things that motivate us, whether monetary or other rewards. In this project, we compared the effect of four mailed messages on activation of high-risk stroke patients: standard, incentive, salience, and incentive plus salience. Our overall aim was to identify which type of stroke MINDSPACE message is most effective in activating persons at high risk of stroke to seek help in developing a personal stroke risk reduction plan. We carried out this project in two health systems that serve a large proportion of individuals that experience disadvantage due to SES, rurality, and race.

## Methods

We conducted this randomized parallel group clinical trial in the Eskenazi Health System (EHS) and the Veterans Health Administration (VA) Indiana System. Both systems have a tertiary medical center facility located next to each other in Indianapolis. EHS is among the five largest safety-net health systems in the U.S. and serves a large proportion of minoritized individuals and individuals with lower SES. In addition to a state-of-the-art hospital, EHS operates 11 Federally Qualified Health Centers throughout Indianapolis, providing more than one million outpatient visits annually. VA Indiana serves more than 62,000 Veterans in Central Indiana with tertiary hospital services in Indianapolis and provides more than 700,000 outpatient visits annually in Indianapolis and 13 other Central Indiana-based clinics. VA Indiana serves a high percentage of individuals with rural home locations and lower SES. This project was approved by the Indiana University Institutional Review Board, which has authority for projects conducted in both health systems, and by the Roudebush VA Research and Development Review committee. The data that support the findings of this study are available from the corresponding author upon reasonable request and to the extent allowable by EHS and VA data use agreements. The study is registered with ClinicalTrials.gov, ID NCT02721446.

Cohort identification: We used electronic health record (EHR) data to construct Framingham Stroke Risk Scores (FSRS) in primary care patients in both healthcare systems. We used previously published algorithms for constructing the FSRS in VA EHR data, including all elements of the FSRS (age; systolic blood pressure at qualifying primary care visit; presence of diabetes, smoking, cardiovascular disease, or atrial fibrillation) except left ventricular hypertrophy, which is not readily available from structured EHR data.^11^ We adapted this methodology to EHS data and used the same ICD-9/10 codes and vital signs data in the Indiana Patient Care data repository and the EHS data repository.^12,13^ Patients were identified as receiving primary care if they had at least one primary care visit with a corresponding blood pressure measurement within the previous 12 months. Patients in the highest quintile of FSRSs were eligible for this project.

Message development: We developed a standard stroke risk message that included the following elements that were common to all four letters: a definition of stroke, a figure depicting an ischemic stroke and occluded blood vessel, and an instruction on how to call a Stroke Prevention Coordinator (Figure 1). The standard letter included only this information, and the incentive letter added a message about receiving a $5 gift card and the entry into a drawing for one of seven $50 gift cards for calling the coordinator. The salience letter included statements designed to make stroke risk more relevant to the patient, including the information that they were at higher risk than others in their health care system, and that “Strokes can cause many different problems including weakness, trouble walking, trouble talking, and in the worst-case inability to take care of yourself.” The incentive plus salience letter included both additional elements. All letters were tested for Flesch-Kincaid readability to increase the likelihood of comprehension among recipients,^14^ and were reviewed by a health communication expert prior to patient testing. We chose a mailed message instead of a text message or other electronic communication based on relatively low prevalence of use of secure communications modalities in both health systems and in order to increase information content of the message.

**Figure 1.**
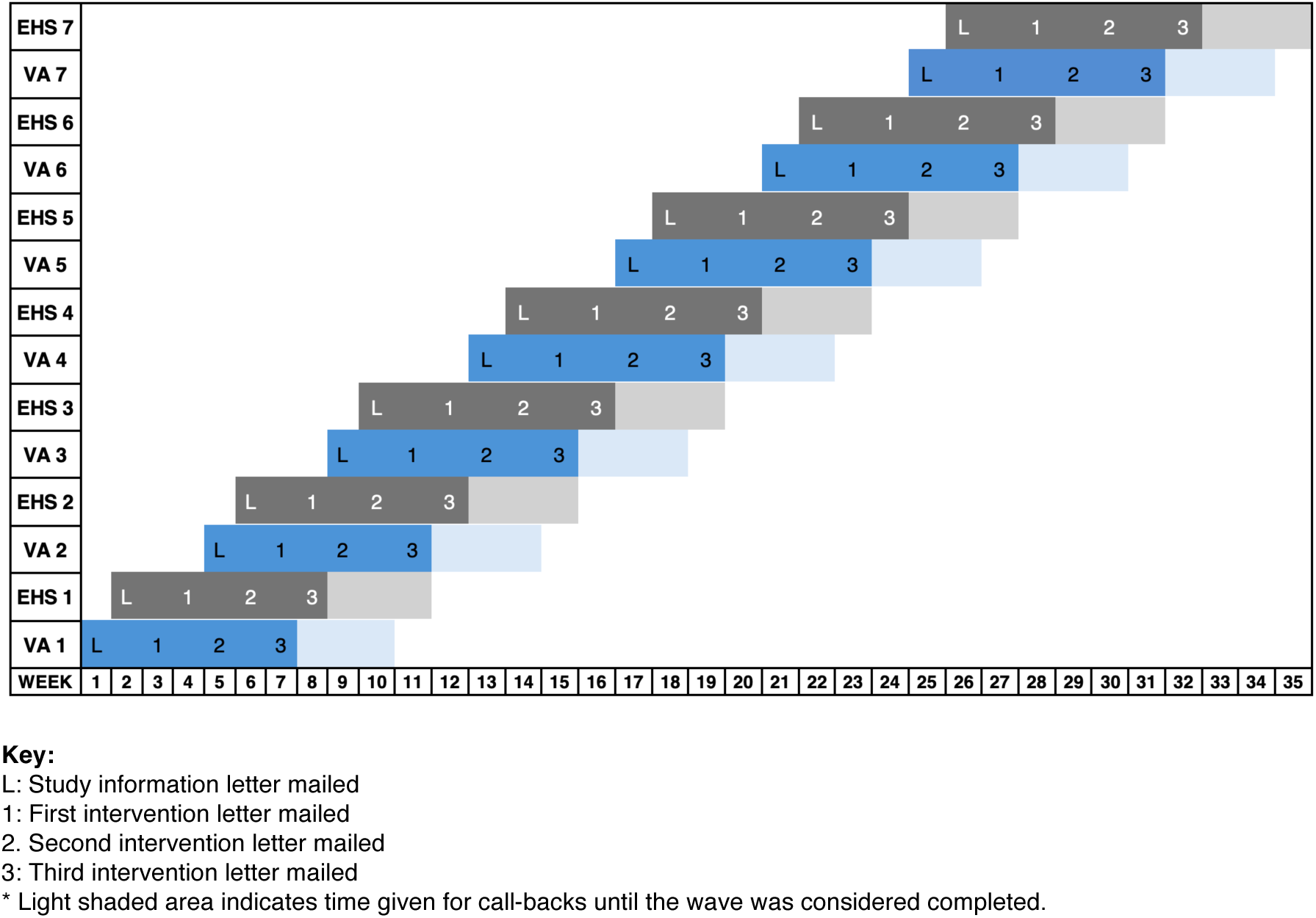
Study mailing timeline Study timeline depicting mailing waves; each wave included a study informational letter (L), followed by up to 3 intervention letters (1, 2, 3) mailed at 2-week intervals.

From the high-risk quintile in both healthcare systems, we recruited 15 individuals to further develop the stroke risk messages. Seven were from EHS, eight from the VA. We conducted in-person interviews about each element of the letter where individuals were encouraged to “think aloud”, a design research strategy that gives valid information about participant thinking.^15^ In addition, each individual was asked to rate the likelihood that they would respond to a given letter on a 10-point scale. Iterative changes to the letters were made throughout this process based on the responses received. Recruitment for the letter development phase ended when information saturation was determined to have occurred.

Intervention: From the highest quantile of the Framingham risk score within each hospital system, for each mailing wave, patients were sampled proportionally from each clinic (after excluding individuals who died or were previously sampled) and randomized 1:1 to the four study arms. EHS had four clinics and VA had two clinics. Randomization was conducted by the study statistician using SAS/STAT^®^ software, version 9.2 of the SAS System for Windows, (SAS Institute Inc., Carey, NC). A random allocation sequence was generated within each clinic and wave. Mailings took place over seven defined waves from August 8, 2016, to March 8, 2017. In each wave, individuals received an informational letter in which the project was described, and they had an opportunity to opt out. If no opt-out request was made within two weeks, the first intervention letter was mailed. If no response was received another letter was sent every two weeks for two more possible mailings (Figure 1). Individuals with an identified bad address were replaced in the next wave with new individuals to attempt to maintain study power. Response to the messages was defined as calling the stroke prevention coordinator.

Patients who called the stroke prevention coordinator completed a brief interview about their awareness of stroke risk factors, health beliefs, readiness to make a change, and what stroke risk factor they would like to work on. The coordinator offered to transfer patients to their primary care provider’s appointment line to make an appointment if that was the desired next step. Patients that left a voicemail message response were called back three times to attempt to complete an interview; those not reached were counted as responding to the mailing.

A waiver of consent and HIPAA was granted to generate individuals’ FSRS and send out mailings. When individuals called the stroke prevention coordinator, they provided an assent to interview.

Statistical analysis: Due to known differences between the two hospital systems in healthcare delivery, availability, and granularity of EHR data as well as age, gender, and race/ethnicity of patients, this study was *a priori* powered to model response to the mailed messages separately in the two cohorts. The study was powered to detect an increase in response (calling the stroke prevention coordinator) of 5% compared to the standard letter in the EHS cohort (estimated 5% to 10%) and an increase of 10% compared to the standard letter in the VA cohort (estimated 30% to 40%). The baseline response rates were estimated from prior studies conducted in these healthcare systems. Assuming the outcome prevalence for the standard message group of EHS individuals is 5%, if we assume the salience message group of EHS is 10%, a Chi-square test with type I error set at .017 (.05/3) for each of the three comparisons will have 80% power to detect this difference with a sample size of 578 per group (total mailed N = 2312). Assuming the outcome prevalence for the standard message group in VA individuals is 30%, if we assume the salience message group for VA is 40%, a Chi-square test with type I error set at .017 (.05/3) for each of the three comparisons will have 80% power to detect this difference with a sample size of 473 per group, (total mailed N = 475 x 4 = 1900). Not all individuals with bad addresses or who had died between randomization and mailing were effectively replaced, however, and thus slightly fewer individuals than planned for received a mailed letter. The final analytic cohort contained N = 2084 EHS and 1759 VA individuals. Within each hospital system cohort, Chi-square and independent t-tests were used to compare demographic and clinical characteristics between those who responded and those who did not respond to the letter.

For each cohort, logistic regression was then used to determine if response to the letter differed by letter type after adjusting for participant demographics, Charlson comorbidity scores,^16^ and FSRS. Profile likelihood confidence intervals are reported for the odds ratios. A Šidák adjustment was used to keep the familywise type I error rate at 5% corresponding to the three pairwise comparisons (with the standard letter as the reference group).^17^ Thus, reported p-values have been adjusted and the type I error is set at .017 for the reported confidence intervals corresponding to the three pairwise comparisons. All statistical analyses were carried out using SAS/STAT software.

## Results

Of 6,695 patients in the highest quintile of stroke risk at participating primary care clinics, 4,422 (2,414 EHS and 2,008 VA) were randomized to receive a letter, 3,843 total individuals received a letter (2,084 EHS and 1,759 VA). Participant characteristics are shown in Table 1. In general, EHS participants were more likely female and Black. VA patients were older, almost exclusively male, and White. Vascular risk factor prevalence, as identified by available inpatient and outpatient ICD-9/10 codes, and the number of outpatient visits in the year prior to study entry were higher in the VA cohort. Accordingly, FSRSs and corresponding mean 10-year stroke risks were also higher in the VA cohort.

**Table 1.**
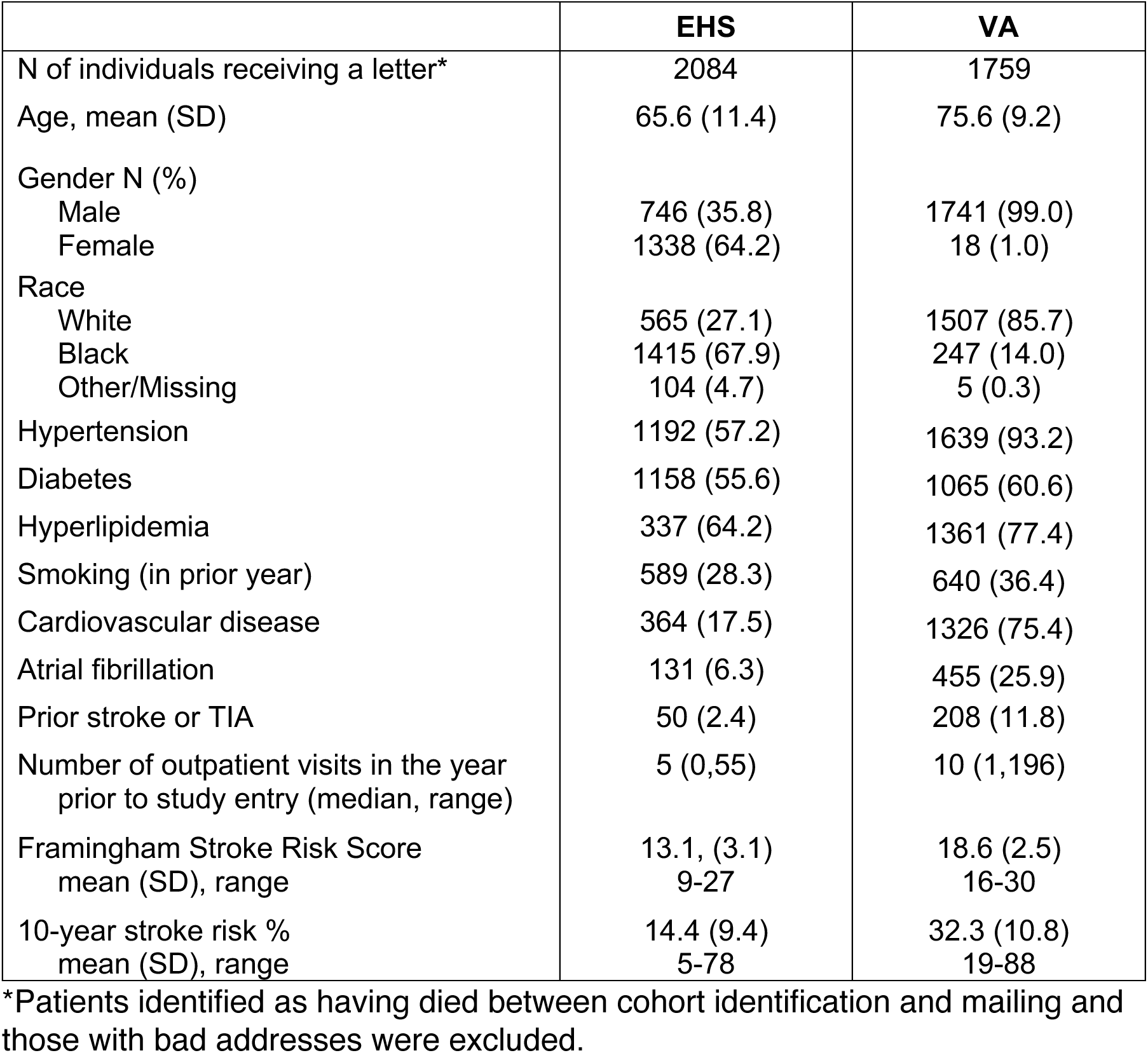
Cohort Demographics.

Overall, 13% of EHS and 23% of VA individuals responded to a letter (Table 2). In both cohorts, the Incentive and the Incentive plus Salience letter generated more responses than the standard letter (Figure 2). For EHS, only the Incentive letter generated statistically significantly more responses than the standard letter, whereas in the VA, only the Incentive plus Salience letter generated significantly more responses. Letters that included a salience message only were not significantly different than the standard letter in either cohort. Call data were obtained for 631 individuals (241 EHS and 390 VA). At least one-quarter of individuals were not aware of their risk factors (31.1% EHS and 26.7% VA), and of those that were aware, only about two-thirds knew that these were stroke risk factors. Most patients (86% EHS and 85% VA) believed they could do things to lower their stroke risk, but fewer were rated as ready to change (defined as being in an action or maintenance stage of change for health behaviors) after completion of the interview (37% in EHS and 44% in VA). EHS patients were more likely to report having made changes in their health for the better in the prior year (68.3% EHS and 52.1% VA).

**Figure 2.**
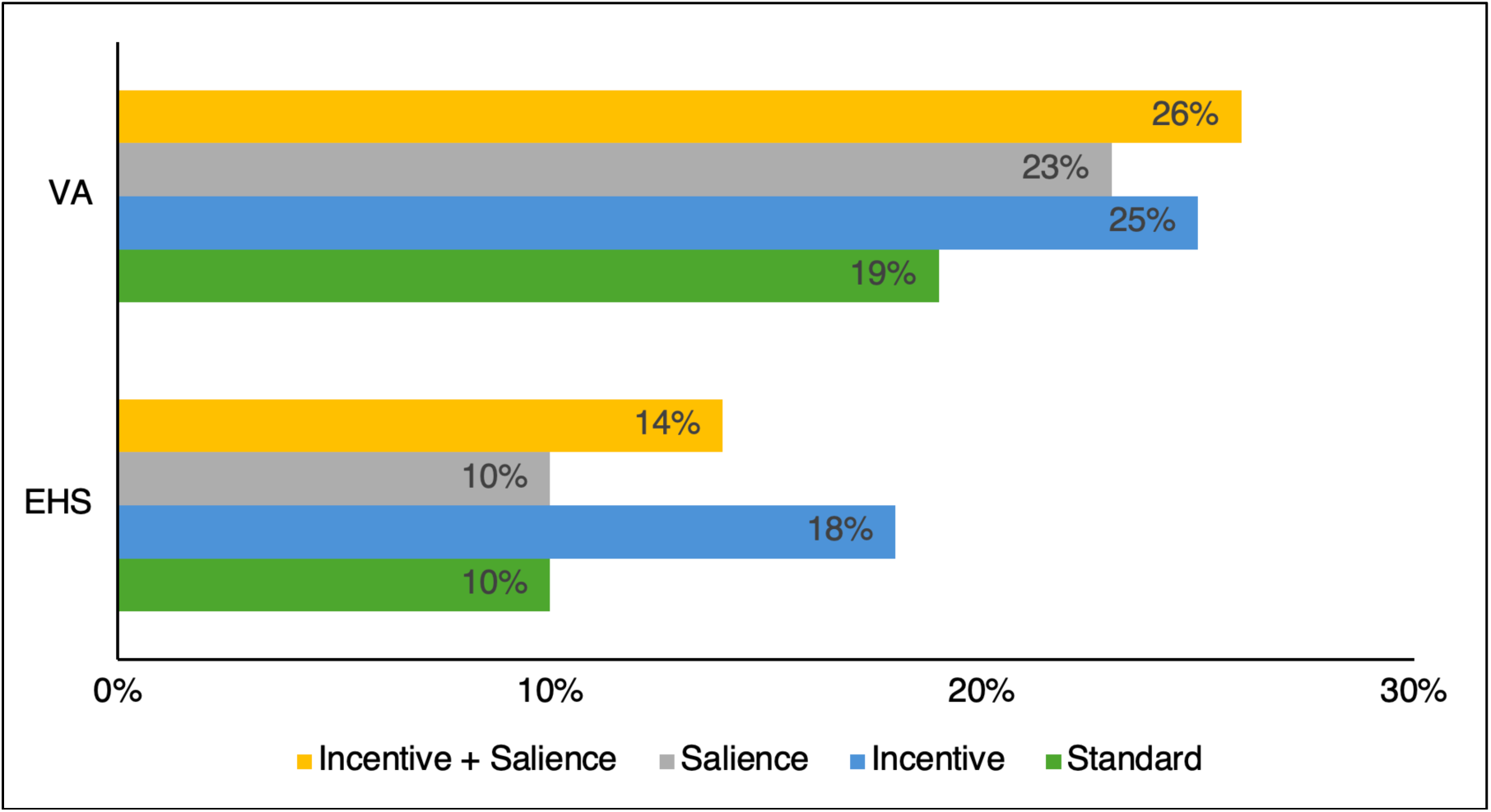
Response to mailed message This figure depicts total responses (calling the stroke prevention coordinator) to each study intervention letter type, by healthcare system (EHS and VA).

**Table 2.** Letter and Call Response Data.

| **Letter Responses\*** | **EHS** | **VA** |
| --- | --- | --- |
| Overall response data | N = 2084 | N = 1759 |
| No response | 1756 (84.3) | 1270 (72.2) |
| Call completed | 241 (11.6) | 390 (22.2) |
| Call but no interview | 32 (1.5) | 19 (1.1) |
| Patient refused | 51 (2.4) | 69 (3.9) |
| Other | 4 (0.2) | 11 (0.6) |
| Response by message group | N = 273 | N = 409 |
| Standard | 53 (19.4) | 86 (21.0) |
| Incentive | 95 (34.8) | 110 (26.9) |
| Salience | 50 (18.3) | 98 (24.0) |
| Incentive plus salience | 75 (27.5) | 115 (28.1) |
| **Call Responses** | **EHS** | **VA** |
| N of individuals that completed call | 241 | 390 |
| Aware of existing stroke risk factors | 166 (68.9) | 286 (73.3) |
| If yes, aware that these are stroke risk factors | 110 (67.5) | 174 (60.8) |
| Belief they can lower their stroke risk | 205 (85.4) | 327 (83.9) |
| Made changes in health in the prior year | 164 (68.3) | 203 (52.1) |
| Ready to change (yes/no) † | 87 (37) | 172 (44) |
| Request to transfer call to PCP | 65 (27.4) | 76 (20.0) |

\*Letter response defined as call completed or call but no interview.
†Ready to change data were available for 238 EHS and 387 VA individuals.

Based on the logistic regression model (Table 3), receiving an incentive-only letter was independently associated with response in the EHS cohort but did not quite reach significance in the VA cohort after adjusting for multiple comparisons (EHS OR 1.97, 95% CI 1.17-3.09, VA OR 1.44, 95% CI 0.98 -2.14). The Incentive plus Salience letter was significantly associated with response in the VA cohort but did not reach significance in the EHS cohort (VA OR 1.50, 95% CI 1.02-2.22, EHS OR = 1.45, 95% CI 0.92-2.30). Demographic factors were variably related to response: in the EHS cohort Black individuals were more than twice as likely to respond than White individuals (OR 2.37, 95% CI 1.73-3.33), and in the VA cohort older age was associated with response (OR 1.02 per year of increased age, 95% CI 1.00-1.03).

**Table 3.** Logistic Regression Results of Letter Response (EHS response = 273/2084, VA response = 409/1759)

|  | **EHS cohort** |  | **VA cohort** |  |
| --- | --- | --- | --- | --- |
|  | OR (95% CI) | p* | OR (95% CI) | p* |
| Incentive vs. Standard | 1.97 (1.17, 3.09) | < 0.001 | 1.44 (0.98, 2.14) | 0.073 |
| Incentive + Salience vs. Standard | 1.45 (0.92, 2.30) | 0.160 | 1.50 (1.02, 2.22) | 0.036 |
| Salience vs. Standard | 0.96 (0.58, 1.58) | 0.996 | 1.25 (0.84, 1.86) | 0.452 |
| Female vs. Male | 0.81 (0.61, 1.07) | 0.139 | --- | --- |
| Age | 1.01 (1.00, 1.03) | 0.174 | 1.02 (1.00, 1.03) | 0.019 |
| Black vs. non-Black | 2.37 (1.73, 3.33) | < 0.001 | 0.98 (0.68, 1.39) | 0.913 |
| Framingham SRS | 0.96 (0.91, 1.02) | 0.160 | 1.00 (0.95, 1.05) | 0.919 |
| Charlson Index | 1.04 (0.95, 1.13) | 0.422 | 0.96 (0.90, 1.02) | 0.200 |

\*Adjusted for multiple comparisons with Šidák adjustment; type I error is set at .017 for the reported confidence intervals.

## Discussion

This study showed that a direct mailed message to high-risk individuals resulted in 13-23% of individuals contacting their healthcare system. Messages that included a $5 monetary incentive for calling were more effective than a standard message, although in the VA population the addition of salience information about how the stroke might affect an individual was associated with the strongest response. Many individuals at high risk of stroke are unaware of their risk, and so may especially benefit from a targeted intervention.

This study is important because data continue to demonstrate that evidence-based interventions to reduce vascular risk are not equally implemented among those at high risk for stroke. For example, Hispanic/Latino/Latina/Latinx adults have lower awareness of hypertension, and both Black and Hispanic/Latino/Latina/Latinx adults have lower blood pressure control compared to White adults.^5,18^ This suggests that strategies like using the FSRS, which includes a measure of blood pressure and not just a diagnosis of hypertension, may more effectively identify patients most likely to benefit from interventions.

Importantly, population-based studies like the Cardiovascular Health Awareness Program demonstrate that increasing awareness of cardiovascular risk and promoting lifestyle and prevention activities and resources can result in sustained improvements in cardiovascular morbidity and mortality.^19^ Specifically in patients with stroke or transient ischemic attack, engaging high-risk individuals in community-based programs may be effective in improving risk factor control and reducing risk of recurrent stroke.^20^ Challenges to enrollment in programs like these include effectively engaging the at-risk population and helping providers identify patients most likely to benefit from program enrollment. Incentives, specifically small monetary incentives, have been shown in some settings to increase patient engagement and to be socially acceptable to patients.^20–22^ The extent to which engagement translates to sustained risk factor modification in high-risk stroke patients is not clear. Nonetheless, this study demonstrates that a risk targeted mailed message with a small incentive may be an effective way to identify and engage a population at high risk of stroke.

We chose to use a mailed message rather than a text message given the age of the group at highest risk for stroke, and the additional message content that a mailed letter allowed us to test over a simple text message. Direct communication to patients via text messaging or EHR-messaging is increasingly used as an intervention to engage patients in improving risk factor control and/or medication adherence but reach among older adults with complex medical conditions is less well known.^23,24^ While older adults are using technology at increasing rates,^25^ use of email and texting platforms decreases with physical capacity limitations.^26^ Recent studies looking at recruitment to risk factor reduction programs demonstrate mail-out recruitment rates of 17-39%,^27,28^ with older age predicting response to mailed letters, which we also observed among the VA cohort.^29^ Moving forward, testing our message on electronic formats is also warranted.

This study has some limitations. Although we enhanced generalizability by including two different health care systems, different availability of data in the EHR made it difficult to directly compare results between the two systems. We also obtained data on diagnoses prior to the index year of study entry, but it is possible that patients with prior stroke are included in the analysis and that this could also impact the likelihood of responding. Additionally, the intervention took place in 2016-2017, however manuscript preparation was halted due to the COVID-19 pandemic leading to delayed publication. As described above, current data still support mailed messages, so we believe these findings remain relevant. Finally, our data do not show whether patient activation in the form of receiving a letter translates to future action, for example having a primary care clinic visit or an explicit discussion about stroke risk with a provider. We are conducting additional analyses with follow-up data to determine whether the messages are related to these more distal actions.

In summary, a simple mailed message with a $5 incentive may be an effective way to increase the engagement of patients at high-risk of stroke with their healthcare system. The addition of salience information may provide additional benefit although patient characteristics most associated with response to salience messages require further investigation. This strategy could serve as a pragmatic means of engaging potentially receptive patients in further evidence-based programs to reduce risk. On a large scale, it is possible that such a strategy could prove cost effective, if the programs offered are successful in promoting and sustaining reductions in vascular risk.

## Data Availability

The data that support the findings of this study are available from the corresponding author upon reasonable request and to the extent allowable by institutional data use agreements.

## Acknowledgements

The authors wish to thank Evgenia Teal from the Regenstrief Institute Data Core for assistance in obtaining EHS system data.

